# Analysis of modular gene co-expression networks reveals molecular pathways underlying Alzheimer’s disease and progressive supranuclear palsy

**DOI:** 10.1101/2021.09.21.21263793

**Authors:** Lukas Iohan Carvalho, Jean-Charles Lambert, Marcos R. Costa

## Abstract

A comprehensive understanding of the pathological mechanisms involved at different stages of neurodegenerative diseases is key for the advance of preventive and disease-modifying treatments. Gene expression alterations in the diseased brain is a potential source of information about biological processes affected by pathology. In this work, we performed a systematic comparison of gene expression alterations in the brains of human patients diagnosed with Alzheimer’s disease (AD) or Progressive Supranuclear Palsy (PSP) and animal models of amyloidopathy and tauopathy. Using a systems biology approach to uncover biological processes associated with gene expression alterations, we could pinpoint processes more strongly associated with tauopathy/PSP and amyloidopathy/AD. We show that gene expression alterations related to immune-inflammatory responses preponderate in younger, whereas those associated to synaptic transmission are mainly observed in older AD patients. In PSP, however, changes associated with immune-inflammatory responses and synaptic transmission overlap. These two different patterns observed in AD and PSP brains are fairly recapitulated in animal models of amyloidopathy and tauopathy, respectively. Moreover, in AD, but not PSP or animal models, gene expression alterations related to RNA splicing are highly prevalent, whereas those associated with myelination are enriched both in AD and PSP, but not in animal models. Finally, we identify 12 AD and 4 PSP genetic risk factors in cell-type specific co-expression modules, thus contributing to unveil the possible role of these genes to pathogenesis. Altogether, this work contributes to unravel the potential biological processes affected by amyloid versus tau pathology and how they could contribute to the pathogenesis of AD and PSP.

## Introduction

Alzheimer’s disease (AD) and progressive supranuclear palsy (PSP) are incurable neurodegenerative disorders that share some common pathological hallmarks, such as synapse loss and the presence of intraneuronal neurofibrillary tangles (NFTs) composed of hyperphosphorylated microtubule-associated protein tau (MAPT or TAU) [1, 2]. AD is also characterized by extracellular amyloid plaques composed of aggregated amyloid-beta peptides and a sustained immune inflammatory response leading to the activation of the brain’s resident macrophages (microglia) and other immune cells [3, 4]. Although these neuropathological features of AD and PSP have been extensively described in *postmortem* brain samples, their precise contribution to pathogenesis remains poorly understood. Recently, RNA-sequencing in large sample cohorts have been used to identify alterations in gene expression associated with onset and progression of AD and PSP [5]. However, no direct comparison of the transcriptional signatures in the brains affected by these two neuropathologies has been performed.

Animal models of tau and amyloid pathology have been largely used to probe AD-related processes [6] and identify gene expression alterations associated with those two different pathological hallmarks [7, 8, 9, 10, 11, 12]. Interestingly, it has been reported that transcriptional perturbations observed in the AD human brain overlap with gene expression alterations observed in the brain of mouse models of AD, frontotemporal dementia, Huntington’s disease, amyotrophic lateral sclerosis and aging [7, 8, 12, 13]. These observations suggest that transcriptional changes in the diseased brain could outline pathophysiological processes and therefore contribute to the understanding of disease mechanisms. However, it remains unclear whether amyloid and tau pathology could lead to similar or distinct transcriptional alterations in the human brain.

In this study, we hypothesized that a systematic analysis of gene expression alterations in brain samples obtained from *postmortem* AD and PSP patients, as well as amyloid and tau pathology animal models, could contribute to disentangle cellular processes affected at different stages of disease progression. To that, we systematically probed gene expression alterations in different brain regions of AD and PSP patients [5], as well as in two different transgenic mouse models used to study amyloid or tau pathology, using different bioinformatics approaches. We also evaluated gene expression at the transcript level, allowing the identification of both differentially expressed genes (DEGs) and genes with different transcript usage (gDTUs) or isoform switches [14].

Our results suggest that inflammatory response is more strongly associated with amyloid pathology in animal models and predominates in the brain of AD patients younger than 80 years. Conversely, synaptic alterations are observed both in young PSP and old AD patients and correlates with tau pathology. Interestingly, we show that isoform switches are abundant in the AD and PSP human brain, but rare in animal models of both amyloid and tau pathology, suggesting that alterations in alternative splicing can be a specific feature of the diseased human brain. Altogether, our work improves our understanding about the biological processes affected by amyloid versus tau pathology and contributes for the development of precise disease-modifying strategies.

## Materials and Methods

### Bulk RNAseq data from human and animal models

All RNAseq datasets used in this work were obtained from AMP-AD Knowledge Portal (https://www.synapse.org) following all terms and conditions to the use of the data. From Mayo Clinic Alzheimer’s Disease Genetics Studies (Mayo), we analyzed RNAseq data generated from two areas (Temporal Cortex and Cerebellum) and 3 types of subjects: individuals with AD; PSP: and elderly individuals with no neurodegenerative disease. To classify the subjects in control (elderly individuals with no neurodegenerative disease) or with one of the two conditions mentioned above we used the ‘Group’ column from metadata obtained from the AMP-AD Knowledge portal (Table 1, Table 2). The “Age_Group” column was used to divide individuals into three groups: A, age of death between 70 and 80 years old; B, age of death between 81 and 89; and C, age of death equal or superior to 90 years old. The number of samples for the Temporal Cortex is: AD - *N = 75* (A = 18, B = 37 and C = 20); PSP - *N = 62* (A = 50, B = 12); Control - *N = 70* (A = 16, B = 34, C = 20). For Cerebellum: AD - *N = 75* (A = 18, B = 37 and C = 20); PSP -*N = 62* (A = 50, B = 12); Control - *N = 70* subjects (A = 16, B = 37, C = 17).

**Table 1:** Summary of clinical and technical variables of samples from MAYO TCX.

| MAYO Temporal Cortex - Human |  |  |  |  |  |  |  |
| --- | --- | --- | --- | --- | --- | --- | --- |
| Age_Group | Sex | n | AOD | Braak | Thal | RIN | PMI |
| AD |  |  |  |  |  |  |  |
| A | female | 10 | 76 (± 3.13) | 5.6 (± 0.52) | 4.8 (± 0.63) | 8.55 (± 0.53) | 5.29 (± 4.82) |
| A | male | 8 | 76.75 (± 2.25) | 5.31 (± 0.46) | - | 8.47 (± 0.38) | 9 (± 8.72) |
| B | female | 23 | 85.13 (± 2.88) | 5.52 (± 0.55) | - | 8.69 (± 0.65) | 6.07 (± 4.57) |
| B | male | 14 | 85.21 (± 2.08) | 5.39 (± 0.63) | - | 8.64 (± 0.65) | 8.09 (± 5.49) |
| C | female | 13 | 90+ | 5.54 (± 0.56) | - | 8.31 (± 0.34) | 8.38 (± 8.93) |
| C | male | 7 | 90+ | 5.29 (± 0.49) | - | 8.63 (± 0.45) | 8.25 (± 5.25) |
| PSP |  |  |  |  |  |  |  |
| A | female | 21 | 75.48 (± 3.76) | 2.1 (± 0.89) | 1.05 (± 1.2) | 8.45 (± 0.47) | 7.78 (± 6.22) |
| A | male | 29 | 75.31 (± 3.06) | 2.14 (± 0.92) | 0.86 (± 1.06) | 8.43 (± 0.53) | 9.27 (± 7.52) |
| B | female | 3 | 82.33 (± 2.31) | 1.17 (± 1.26) | 0.33 (± 0.58) | 8.23 (± 0.25) | 11.33 (± 2.08) |
| B | male | 9 | 84.11 (± 2.37) | 2.39 (± 0.99) | 1.11 (± 1.17) | 8.69 (± 0.53) | 3.5 (± 0.71) |
| Control |  |  |  |  |  |  |  |
| A | female | 3 | 76.67 (± 2.08) | - | - | 7.77 (± 1.54) | 3 (± 1.41) |
| A | male | 13 | 76 (± 2.97) | - | - | 7.25 (± 0.95) | 2.6 (± 0.84) |
| B | female | 19 | 86.47 (± 2.04) | - | - | 7.93 (± 0.81) | 4.5 (± 3.14) |
| B | male | 15 | 84.87 (± 2.9) | - | - | 7.68 (± 0.83) | 5.71 (± 5.55) |
| C | female | 12 | 90+ | - | - | 7.06 (± 0.92) | 13.78 (± 10.96) |
| C | male | 8 | 90+ | 2.31 (± 1) | - | 8.28 (± 1.51) | 11.29 (± 10.21) |

**AOD** = Age of Death, **n** = number of samples, **RIN** = RNA integrity number, **PMI** = Postmortem interval (in hours).
**Braak** = Neurofibrillary tangles staging, **Thal** = Thal amyloid staging.
**AD** = Alzheimer's Disease.
**PSP** = Progressive Supranuclear Palsy. Values are mean ± SD.

**Table 2:** Summary of clinical and technical variables of samples from MAYO CER.

| MAYO Cerebellum - Human |  |  |  |  |  |  |  |
| --- | --- | --- | --- | --- | --- | --- | --- |
| Age_Group | Sex | n | AOD | Braak | Thal | RIN | PMI |
| AD |  |  |  |  |  |  |  |
| A | female | 10 | 76 (± 3.13) | 5.6 (± 0.52) | 4.8 (± 0.63) | 8.39 (± 0.86) | 5.29 (± 4.82) |
| A | male | 8 | 76.75 (± 2.25) | 5.31 (± 0.46) | - | 7.84 (± 1.15) | 9 (± 8.72) |
| B | female | 22 | 84.91 (± 2.76) | 5.59 (± 0.53) | - | 8.46 (± 0.72) | 4.86 (± 3.03) |
| B | male | 15 | 85.13 (± 2.03) | 5.43 (± 0.62) | - | 8.24 (± 0.9) | 7.82 (± 5.62) |
| C | female | 13 | 90+ | 5.42 (± 0.53) | - | 8.41 (± 0.27) | 15.71 (± 13.59) |
| C | male | 7 | 90+ | 5.29 (± 0.49) | - | 8.13 (± 0.59) | 11.5 (± 7.68) |
| PSP |  |  |  |  |  |  |  |
| A | female | 21 | 75.48 (± 3.76) | 2.1 (± 0.89) | 1.05 (± 1.2) | 8.68 (± 0.91) | 7.78 (± 6.22) |
| A | male | 29 | 75.28 (± 3.06) | 2.12 (± 0.91) | 0.93 (± 1.07) | 8.22 (± 0.97) | 8.75 (± 7.4) |
| B | female | 3 | 82.33 (± 2.31) | 1.17 (± 1.26) | 0.33 (± 0.58) | 8.07 (± 0.97) | 11.33 (± 2.08) |
| B | male | 9 | 84.11 (± 2.37) | 2.39 (± 0.99) | 1.11 (± 1.17) | 8.3 (± 0.9) | 3.5 (± 0.71) |
| Control |  |  |  |  |  |  |  |
| A | female | 3 | 76.67 (± 2.08) | - | - | 7.5 (± 1.05) | 2.67 (± 1.15) |
| A | male | 13 | 76.15 (± 2.85) | - | - | 7.57 (± 1.18) | 3.75 (± 3.93) |
| B | female | 21 | 85.86 (± 2.2) | - | - | 7.54 (± 1.02) | 4.24 (± 3.01) |
| B | male | 16 | 85.12 (± 2.99) | - | - | 7.64 (± 0.94) | 5.71 (± 5.55) |
| C | female | 10 | 90+ | - | - | 7.1 (± 1.02) | 13.89 (± 11.33) |
| C | male | 7 | 90+ | - | - | 8.49 (± 0.64) | 13.8 (± 11.32) |

**AOD** = Age of Death, **n** = number of samples, **RIN** = RNA integrity number, **PMI** = Postmortem interval (in hours).
**Braak** = Neurofibrillary tangles staging, **Thal** = Thal amyloid staging.
**AD** = Alzheimer's Disease.
**PSP** = Progressive Supranuclear Palsy.
Values are mean ± SD.

We used two animal models in this study: 5XFAD and TauD35. In the 5XFAD [37] model we analyzed RNAseq data from hippocampus. We classified the animals into two groups, “Alzheimer” and “Control”, and subdivided them using the column “Age_Group” in three groups (4 M, 12 M, and 18 M, where M represents the month of the death), which can be found in the metadata from AMP-AD Knowledge portal (Table 4). Thereby, in the Alzheimer’s group we have 10 animals from subgroup 4 M, 9 from subgroup 12 M, and 16 from subgroup 18 M. In the control group, we have 10 animals from subgroup 4M, 10 animals from subgroup 12 M, and 20 from subgroup 18M. The TauD35 [38] model has RNAseq data from the hippocampus. Like the 5XFAD model, we classified the animals in “Alzheimer” and “Control” groups, subdividing them into 2 subgroups according to the month of the death of the animal. Alzheimer’s groups have 9 animals, subdivided in 4 M (5 subjects) and 17 M (4 subjects); control groups have 11 animals, subdivided in 4 M (5 subjects) and 17 M (6 subjects).

**Table 3:**
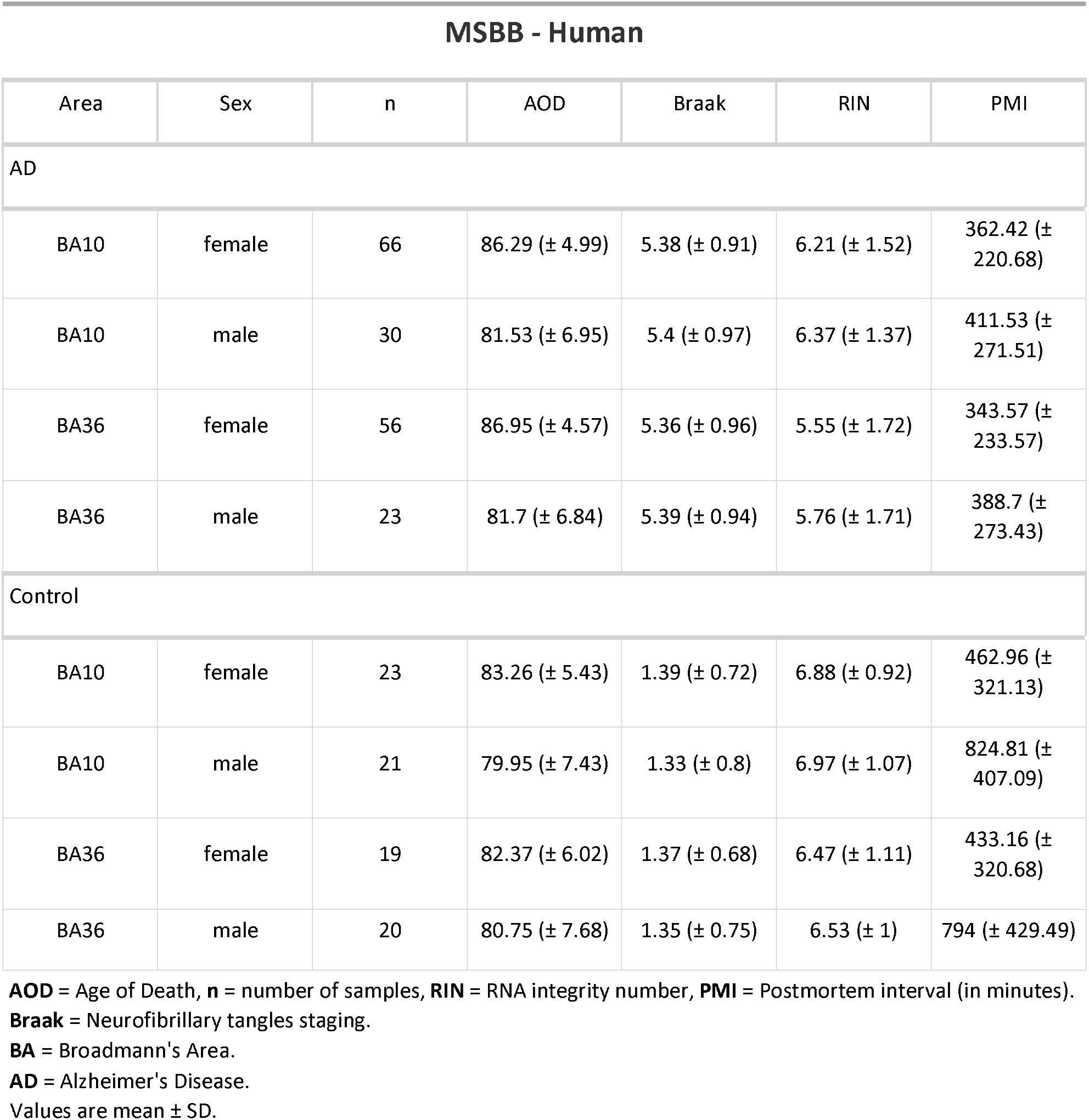
Summary of clinical and technical variables of samples from MSBB.

**Table 4:** Summary of age, sex and technical variables of samples from Animal Models.

| 5XFAD-Taud35 Hippocampus - Animal Model |  |  |  |  |
| --- | --- | --- | --- | --- |
| Age_Group | Sex | n | Group | RIN |
| 5XFAD |  |  |  |  |
| 4 M | female | 10 | Control | 9.58 (± 0.26) |
| 4 M | female | 10 | Alzheimer | 9.73 (± 0.19) |
| 4 M | male | 10 | Control | 9.09 (± 0.73) |
| 4 M | male | 10 | Alzheimer | 9.62 (± 0.27) |
| 12 M | female | 8 | Control | 9.89 (± 0.1) |
| 12 M | female | 10 | Alzheimer | 9.73 (± 0.21) |
| 12 M | male | 12 | Control | - |
| 12 M | male | 8 | Alzheimer | 9.89 (± 0.08) |
| 18 M | female | 32 | Control | - |
| 18 M | female | 12 | Alzheimer | - |
| 18 M | male | 20 | Alzheimer | - |
| 18 M | male | 8 | Control | - |
| TauD35 |  |  |  |  |
| 4 M | female | 1 | Alzheimer | 8.9 |
| 4 M | male | 4 | Alzheimer | 8.7 (± 0.29) |
| 4 M | male | 4 | Control | 8.72 (± 0.15) |
| 4 M | female | 1 | Control | 8.9 |
| 17 M | male | 3 | Alzheimer | 8.47 (± 0.23) |
| 17 M | female | 1 | Alzheimer | 8.8 |
| 17 M | female | 4 | Control | 8.57 (± 0.3) |
| 17 M | male | 2 | Control | 8.55 (± 0.49) |

**M** = Months, **n** = number of samples, **RIN** = RNA integrity number. Values are mean ± SD.

To evaluate transcription signatures in different brain regions of the same patients, we used RNAseq data from the Mount Sinai/JJ Peters VA Medical Center Brain Bank (MSBB–Mount Sinai NIH Neurobiobank) cohort (MSBB), which can be found in the metadata from AMP-AD Knowledge portal (Table 3). We analyzed gene expression alterations in two brain areas known to be affected at early and late stages of AD pathology, respectively: lateral perirhinal cortex (PRh, BA 36) and rostrolateral prefrontal cortex (RLPFC, BA 10). Sample sizes are: PRh - *N = 172* (AD = 128, Control = 44); RLPFC - *N = 174* (AD = 121, Control = 53).

### Realignment of human and animal models reads with Kallisto

We used the pseudoaligner tool Kallisto (version 0.41.1) [39] to align all fastq files obtained from AMP-AD Knowledge Portal (Fig 1). The reference used in the first step of the pipeline to build an index was GRCh38 cDNA release 94 (http://ftp.ensembl.org/pub/release-94/fasta/homo_sapiens/cdna) for the human data and the GRCm38 cDNA release 94 (http://ftp.ensembl.org/pub/release-94/fasta/mus_musculus/cdna) for both mouse animal models.

**Fig 1:**
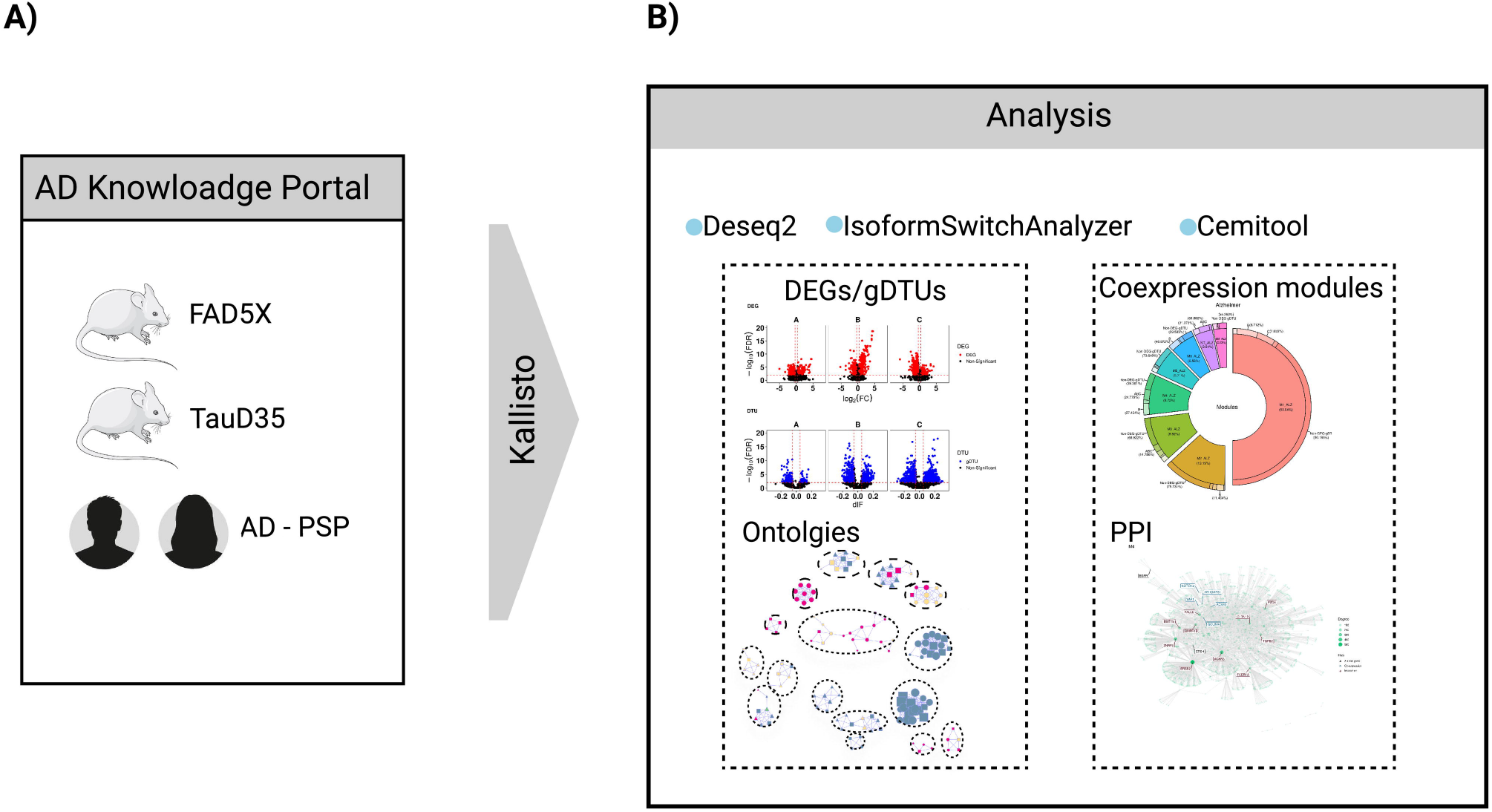
Schematic summary of methodology. A) Human (AD, PSP) and mouse (5XFAD, TauD35) RNA-seq data were obtained from the AD Knowledge Portal and grouped according to the age of death. Next, RNAseq data was pseudo-aligned using Kallisto. B) Analyses were performed using four R packages: DESeq2, IsoformSwitchAnalyzer, gprofiler2 and CEMiTool.

### Differential gene expression analysis

To discover differentially expressed genes (DEGs) we used the R library DESeq2 [40] with the gene expression at transcription-level strategy, as described in our last work [14]. Kallisto output was import in R (version 4.0.5) programming environment and a DESeq2 object was created and filtered based on sum of rows with counts bigger than 10. Next, *DESeq* function was used to filter results based on a model using the negative binomial distribution and correction method FDR (False Discovery Rate). Genes were considered differentially expressed only with FDR < 0.01 and absolute fold change (|FC|) > 1.3.

Isoform switch/differential transcript usage (DTU) analysis was performed using the R library IsoformSwitchAnalyzeR [41]. The analysis was made following the pipeline instructions of the package. First, we imported kallisto abundance tables (same used in DESeq2 pipeline) using *importIsoformExpression* and *importRData* functions to create a *switchAnalyzeRlist* object. Equal to kallisto realignment we used the same cDNA release and its correspondent annotation (http://ftp.ensembl.org/pub/release-94/gtf/homo_sapiens/Homo_sapiens.GRCh38.94.chr_patch_hapl_scaff.gtf.gz) as input. Data was filtered using gene expression cutoff = 3, differential isoform fraction (dIF) = 0.01 and removed single isoform genes. Results of this first part were used in the second part of the analysis with addition of external analysis (CPC2, Pfam, SignalIP and Netsurfp2), as indicated in the pipeline of IsoformSwitchAnalyzer. To confirm our results, we used the R package stageR [42] to generate isoforms overall false discovery rate (OFDR). We selected only isoforms with OFDR < 0.01 and |dIF| > 0.01. Similar parameters were used for analyses of RNA-seq data from animal models, except for the use of GRCM38 cDNA release 94, as mentioned above for pseudoalignment (http://ftp.ensembl.org/pub/release-94/gtf/mus_musculus/Mus_musculus.GRCm38.94.chr_patch_hapl_scaff.gtf.gz).

### Gene set enrichment analysis (GSEA) and Gene Ontology Network

For the gene ontology analysis, we used the R library gprofiler2 [43]. In the function *gost*, we set the parameters correction_method = “FDR” and significant=TRUE and a set of genes, divided into 3 groups: DEGs, gDTUs, and DEGs-gDTUs. We did this to all conditions and groups based on the age of death (A, B, and C for human data; 4M, 12M, 17M, and 18M for mouse data). The filters in this analysis were: false discovery rate (FDR) < 0.01; intersection size (intersection between gene set vs. a few genes in a term) > 3; and precision (intersection size divided by gene set) >0.03. Gprofiler2 has its own default method for computing multiple testing correction for p-values gained from G0 and pathway enrichment analysis. As described by the authors of the package, it corresponds to an experiment-wide threshold of a 0.05, i.e., at least 95% of matches above threshold area statistically significant. We used the Gene Ontology (GO or by branch GO:MF, GO: BP, GO:CC) category to create the table with the results. For the construction of the gene ontology network, we used the results as Gene Matrix Transposed files (gmt): gmt files are archives with gene ontology terms and those genes enriched to the terms. To identify enriched terms sharing the same genes, we used the gmt file containing all gene ontology terms and the associated genes from the human species provided in gprofiler2’s website (gprofiler_full_hsapiens.name.gmt - https://biit.cs.ut.ee/gprofiler/static/gprofiler_hsapiens.name.zip).

We import this information in Cytoscape version (3.9.0) [44] and use the gmt expression files for each group, as mentioned before, and the gmt annotation file to construct the network of ontologies with the Enrinchmap plugin (version 3.3.3) [45]. As result, we had a network of gene ontology terms where nodes correspond to gene ontology terms and edges to terms sharing the same genes. The node and edge tables from networks were exported and imported into R for a better visualization using the R library RedeR [46]. Only interactions with FDR < 0.05 and nodes with degree higher than 2 were shown. The name of a group of nodes was selected by the node with the higher degree.

### Gene co-expression and module analysis

We used the R library package CEMiTool [19] to identify and analyze gene co-expression modules. CEMiTool uses a gene expression file containing genes as rows and samples as columns; we used the matrix create in DESeq2 analysis. This matrix is filtered by an unsupervised method based on the inverse gamma distribution, which will then select the genes used in the analyses. Similarity between pair of genes is evaluated with a modified algorithm created by the authors of the package and then genes are grouped into modules using the Dynamic Tree Cut package [47]. To know which and how many modules in each condition had, we used the main function *cemitool*. For the arguments, we used the normalized expression matrix of counts, metadata from every condition, *p_value* <0.1 (as suggested by the authors of the study), and *ora_pval < 0*.*01*. In this analysis, we did not subdivide subjects according to the age of death, just by condition, i.e, AD, PSP, and control for human data and Alzheimer and control for mouse data. The final object from the analysis has some information about the identified modules: which genes belong to the modules, Over Representation Analysis (ORA), Normalized Enrichment Score (NES), and Protein Protein Interaction (PPI). All these results were retrieved with the function *write_files()*. In ORA plots we showed only the GO terms with FDR<0.01. To identify which modules are up or down-regulated between conditions, CEMiTool uses the fgsea [23] package. If this enrichment is significant, this information is summarized in the variable (NES), which is the enrichment score for a module in each class normalized by the number of genes in the module. Plots showing NES are only those modules with FDR<0.01.

The PPI networks in the Fig 7 show networks with AD genes risk and PSP gene risk. The PPI of these figures were retrieved from the final object of CEMiTool analysis and contains interactions between proteins identified in modules. It is important to note that CEMiTool uses an interaction file (PPI) to construct the interaction network. In this work we decided to show only modules with AD or PSP risk genes.

### Analysis of gene set signatures enriched in unique neural cell types using Cell-ID

To evaluate the enrichment of module-specific gene set signatures in single-cell types, we used the R package Cell-ID [21]. This package allows a clustering-free multivariate statistical method for the robust extraction of per-cell gene signatures from single-cell RNASeq. In this work we used the single-cell signature from Leng’s dataset [22], and performed multiple correspondence analysis (MCA) in scRNA-seq data from the entorhinal cortex of 3 healthy (Braak 0) individuals. Next, with CelI-ID we extracted and calculate the enrichment of per-cell gene signatures using as reference the list of genes from each module (S4 table) identified by CEMiTool. Statistical significance of this enrichment is calculated using a hypergeometric test and shown in function of - log10(pvalue).

## Results

### Gene expression alterations in the neocortex of AD and PSP patients

PSP is a primary tauopathy with abnormal accumulation of tau protein within neurons as neurofibrillary tangles (NFTs), primarily in the basal ganglia, diencephalon, brainstem, and cerebellum, with restricted involvement of the neocortex [29]. On the other hand, AD can be considered as a secondary tauopathy, since Aβ plaques are closely tied to the primary neuropathological process, with primary involvement of transentorhinal region and entorhinal cortex [25]. To investigate the similarities and differences in gene expression alterations in the brain of patients with AD, PSP and elderly controls without clinic-pathological signs of neurodegenerative diseases. These data are available from the MayoRNAseq study, with whole transcriptome data for 275 Cerebellum (CBE) and 276 Temporal cortex (TCX) samples from 312 North American Caucasian subjects with neuropathological diagnosis of AD, PSP, pathologic aging (PA) or elderly controls (CON) without neurodegenerative diseases [5]. We subdivided samples per age (Table 1, Table 2), which is the only metadata common to the different groups of patients and strongly correlates with pathological progression both in AD and PSP [15, 16, 17]. We observed that both the number of differentially expressed genes (DEGs) and genes with differential transcript usage (gDTUs) detected in the TCX of AD patients compared to controls increased with age (Fig 2). Notably, there was very little overlap in the DEGs and gDTUs observed at different ages, suggesting that singular gene expression alterations prevail in the AD brain at distinct pathological stages (Fig S1, Table S1). Accordingly, gene set enrichment analysis (GSEA) revealed that DEGs and gDTUs identified in the AD TCX were significantly enriched for distinct gene ontologies (GOs) according to age (Fig 2A, Table S2). While significant enrichment for GOs associated with immune-inflammatory response, RNA splicing, BMP signaling pathway and gliogenesis were already observed in group A, B and C, terms associated with ion homeostasis, Wnt signaling pathway, cellular response to lipid were exclusively detected in group B. The terms cellular respiration, protein targeting to ER and regulation of protein catabolic processes were solely observed in group C, whereas terms related to synapse signaling were mainly observed in groups B and C. As previously reported, enrichment for the latter terms was only observed when inputting gDTUs alone or in combination with DEGs (Fig 2A), supporting the view that isoform-switches are an important source of gene expression alterations affecting synapses [14]. Analyses of gene expression alterations in the cerebellum of the same patients also revealed a weak overlap among DEGs and gDTUs observed at different ages (Fig S1). Yet, DEGs and gDTUs in all groups were enriched for GOs associated with regulation of neuron projection development, synapse signaling, mRNA metabolic processes and RNA splicing (Fig S2), as observed in the TCX (Fig 2A). These observations suggest that some biological processes are commonly altered in the TCX and cerebellum of AD patients, whereas others, such as immune-inflammatory response, ion homeostasis, BMP and Wnt signaling pathways are mainly affected in the TCX.

**Fig 2:**
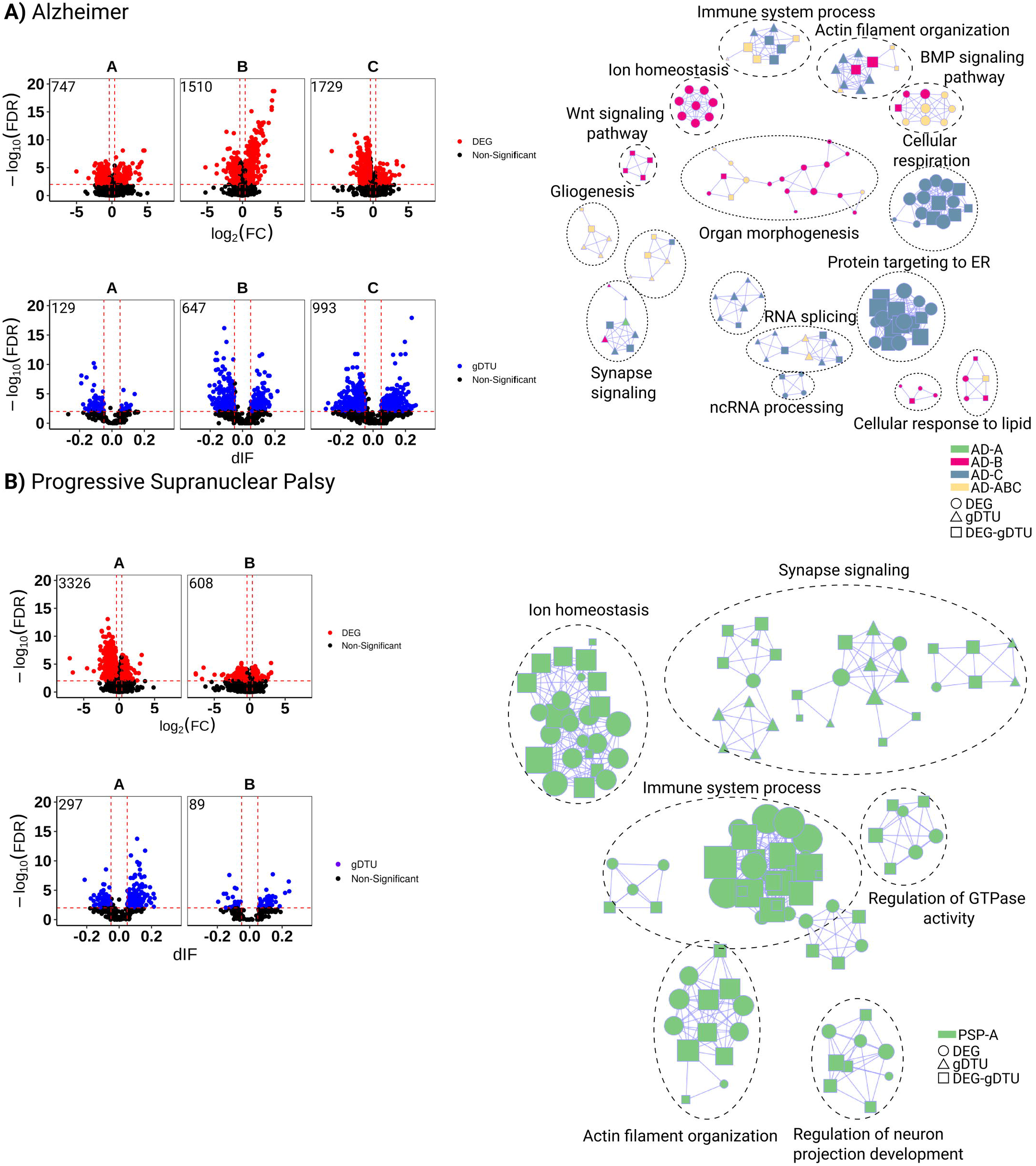
Gene expression alterations in the temporal cortex of AD and PSP patients. A) Volcano plots (left) showing differentially expressed genes (DEGs, red dots; FC > 1.3 and FDR < 0.01), genes with differential transcript usage (gDTU, blue dots; Differential isoform fraction (dIF) and FDR < 0.01) and a network representation (right) of gene ontologies (GOs) significantly enriched in AD. Circles, triangles and squares indicate, respectively, GOs enriched for DEGs, gDTUs or a combination of both. Colors in the network indicate groups where gene expression alterations were detected. B) Same for PSP. Numbers of significantly altered genes identified in each analysis are shown in the top left corner of the volcano plots.

In contrast with AD brains, where few DEGs could be detected at early ages, we found a high number of DEGs and gDTUs in the TCX of PSP patients in group A, consistent with the fast progression of this disease [18]. In this group, DEGs and gDTUs were significantly enriched for several GOs observed in AD patients including those associated with synapse signaling, immune system process, ion homeostasis and regulation of neuron projection development (Fig 2B, Table S2). The number of DEGs and gDTUs identified in elderly PSP patients (group B) was significantly lower than in group A and did not show any enrichment for GOs (Fig 2B). This could be due to the reduced number of samples in group B or to a lower pathological burden in the neocortex of these PSP patients, reflecting the clinic-pathological heterogeneity of PSP [19]. According to this second possibility, we observed an inverted pattern in the distribution of DEGs and gDTUs in the cerebellum of PSP patients, namely a higher number of altered genes in group B (Fig S1). However, only DEGs/gDTUs from group A showed significant enrichment in GSEA (Fig S2B). Some enriched GOs observed in the cerebellum were also detected in the TCX, such as ion homeostasis and regulation of neuron projection development but did not show any enrichment for terms related to immune system processes (Fig S2A). Together with our observations in AD patients, these observations suggest that gene expression alterations associated with immune-inflammatory response are mainly restricted to the neocortex of both AD and PSP patients.

### Gene network analyses reveal cell-type specific molecular pathways in AD and PSP

To further exploit transcriptomic data obtained from in AD and PSP patients and uncover the latent systems-level functionality of genes, we analyzed modular gene co-expression networks using CEMiTool [20]. We detected eight different modules in the TCX of AD and PSP individuals (Fig 3A). Modules 1 and 2 in AD (M1_AD and M2_AD) and modules 1 and 3 in PSP (M1_PSP and M3_PSP) were significantly enriched for genes associated with synaptic signaling (Fig3A, Table S3). M3_AD and M2_PSP were significantly enriched for myelination, whereas M5_AD, M8_AD, M4_PSP and M8_PSP were significantly enriched for several terms associated with immune-inflammatory processes. The modules M6_AD, M7_AD and M6_PSP were associated with extracellular matrix and cell differentiation/angiogenesis, whereas M4_AD and M5_PSP were associated with cellular response to growth factors. In PSP, we also detected a module (M7) associated with apoptosis (Fig3A, Table S3).

**Fig 3:**
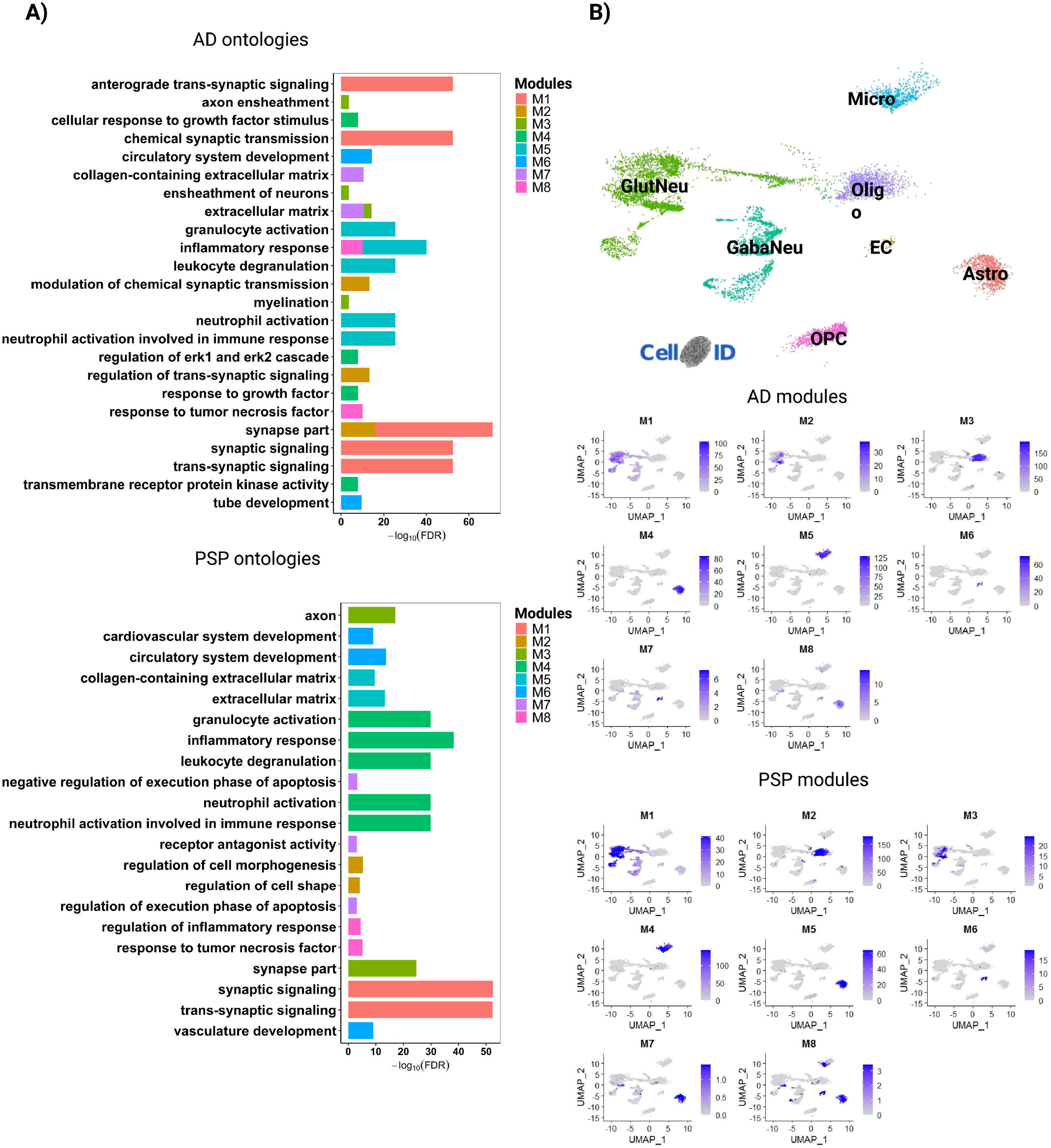
Gene co-expression modules are associated with cell-type specific processes. A) Over Representation Analysis (ORA) for modules identified in AD and PSP in temporal cortex RNA-seq data. Only ontologies with FDR < 0.01 are shown.B) Dimension plot showing major cell types identified in a scRNA-seq dataset generated from the adult human brain [22]. C) Feature plots showing the enrichment of genes identified in different modules. Scale indicates - log10(Pvalue) for the hypergeometric test used in CellID. Astro (Astrocyte), EC (Endothelial cell), GabaNeu (Gabaergic Neuron), GlutNeu (Glutamatergic Neuron), Micro (Microglia), Oligo (Oligodendrocyte), OPC (Oligodendrocyte Precursor Cell).

Next, we used Cell-ID [21] to categorize the cell-types of the adult human brain enriched for the gene signatures identified in the different modules. To that, we performed multiple correspondence analysis (MCA) in single-cell RNA-sequencing (scRNA-seq) data obtained from the entorhinal cortex of three AD patients at early stages of the pathology (Braak 0) [22] (Fig 3B). Then, we calculated the enrichment of per-cell gene signatures using as reference (i) the list of genes identified in each module (Table S4) and (ii) the per-cell gene signatures extracted through Cell-ID from the scRNA-seq data. We observed that modules associated with synapses were mainly enriched in glutamatergic neurons, whereas those associated with myelination were consistently enriched in oligodendrocytes (Fig 3A, Fig 3C). M5_AD and M4_PSP were exclusively enriched in microglial cells, consistent with their enrichment for immune-inflammatory processes. However, M8 in both AD and PSP, which was also associated with inflammation, showed a significant enrichment in astrocytes. These cells were also significantly enriched for M4_AD and M5_PSP (cellular response to growth factors) and M7_PSP (apoptosis). Finally, we found a significant enrichment of M6_AD, M7_AD and M6_PSP gene signatures in endothelial cells.

### Activity of modules is differently altered in AD and PSP brains

Using the fgsea (Fast Gene Set Enrichment Analysis) package [23] built-in CEMitool, we analyzed the association of module activity to sample phenotypes. In this analysis, genes from co-expression modules are treated as gene sets and the z-score normalized expression of the samples within each phenotype is ranked, providing an assessment of modules across different phenotypes [20]. We observed that the normalized enrichment score (NES) of M4, M6, M7 and M8 was greatly higher in AD patients, whereas activity of M1 and M3 was higher in PSP patients compared to control subjects (Fig 4A). Conversely, NES of M2, M4, M6, M7 and M8 in PSP and M1, M2 and M3 in AD brains was marginally lower than in controls (Fig 4A). These findings suggest that gene expression alterations associated with the immune-inflammatory system, cellular response to growth factors, extracellular matrix and cell differentiation/angiogenesis predominate in AD, whereas those associated with synapses and myelination prevail in PSP. According to this interpretation, quantification of the proportion of genes with altered expression within modules revealed that DEGs/gDTUs were more numerous in synaptic modules of PSP than in AD (Fig 4B). Interestingly, genes with altered expression in these synaptic modules were mainly detected at older ages (groups B and C) in AD brains (Fig 4). Conversely, the frequency of DEGs/gDTUs in modules associated with extracellular matrix, cell differentiation/angiogenesis and response to growth factors was much higher in AD than in PSP brains. The modules associated with myelination and immune-inflammatory responses showed a high proportion of DEGs/gDTUs in both diseases and they were highly frequent at early ages (Fig 4B, Fig 4C).

**Fig 4:**
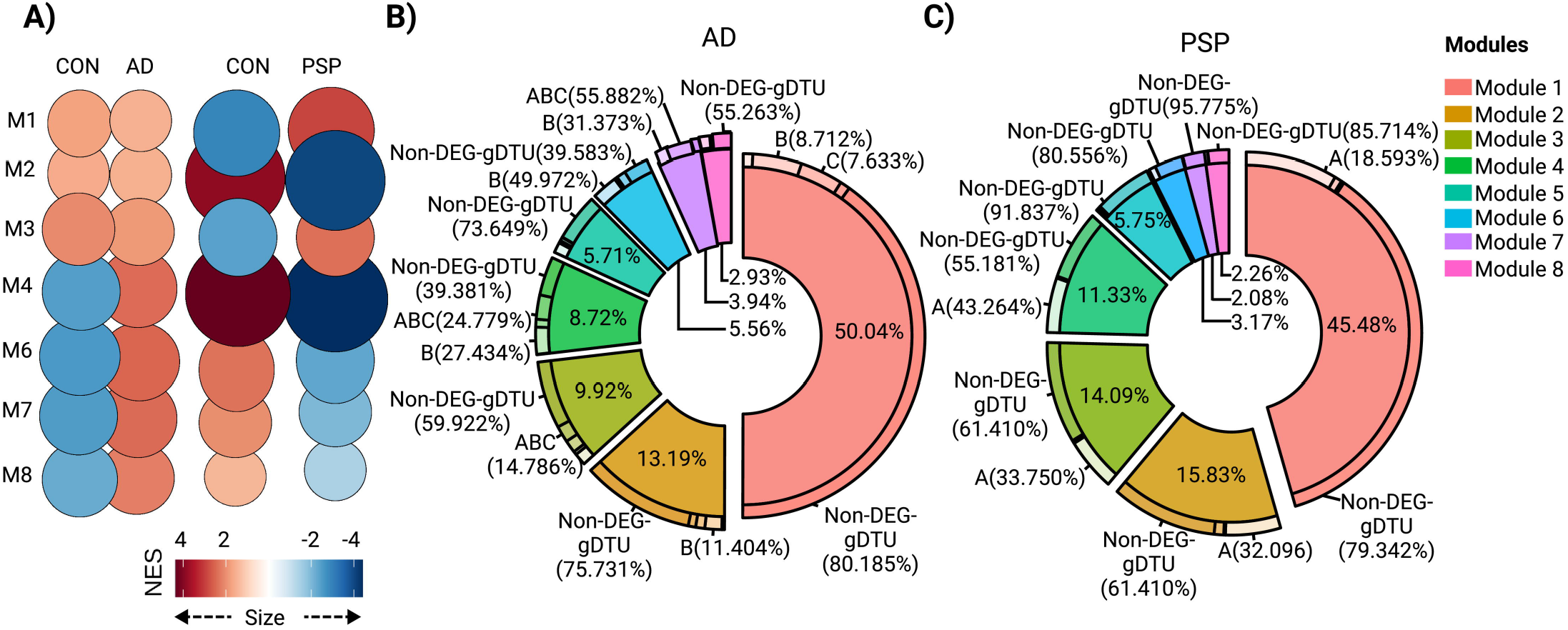
Module enrichment in AD and PSP indicate different alterations in synaptic transmission and immune-inflammatory response. A) Normalized enrichment score (NES) for modules identified in AD, PSP and control from temporal cortex data. Size of circles is equal to the absolute value of NES. Color of circles represents up (red) or down (blue) regulation between diseases (AD or PSP) versus control and is proportional to NES value. Only modules whose comparison reached statistical significance (FDR<0.01) are shown. B) Quantification of the proportion of genes contributing to each module and the percentages of genes within modules that were DEGs or gDTUs in different groups to AD. C) Same for PSP.

To directly probe a possible link between pathology progression and gene expression alterations in the AD brain, we analyzed RNAseq data from the Mount Sinai/JJ Peters VA Medical Center Brain Bank (MSBB–Mount Sinai NIH Neurobiobank) cohort, which encompassed transcriptome sequencing data from multiple brain regions of 364 postmortem control, mild cognitive impaired (MCI) and AD brains with rich clinical and pathophysiological data [24]. Using this dataset to compare transcriptional signatures of different brain regions of AD patients, we have previously shown that gene expression alterations are more prominent in the Broadmann area 36 (perirhinal cortex – PRh) than in the Broadmann area 10 (rostrolateral prefrontal cortex - RLPFC) [14], thus providing an interesting source to observe transcriptional changes associated with pathological progression [25]. Using CEMItool, we identified 4 gene co-expression modules both in RLPFC and PRh (Fig S3A, Table S3). Interestingly, activity of M2 in the RLPFC (affected at late pathological stages) was higher in AD patients and genes in this module significantly enriched for GOs associated with immune-inflammatory responses (Fig S3B). In this same brain region, no modules associated with synapses could be identified (Table S3). Conversely, M1 and M2 in PRh (affected at early pathological stages) significantly enriched for several synapse related GOs and showed a weaker activity in AD patients compared to controls (Fig S3C, Table S3). Collectively, these observations in two independent datasets suggest that gene expression alterations related to immune-inflammatory processes are an early event in the pathogenesis of AD, whereas gene expression changes related to synapses are mainly a late outcome.

### Progressive gene expression alterations in 5XFAD and TauD35 mice

To further investigate the correlation between pathological progress and gene expression alterations, we took advantage of RNA-seq data generated from the brain of animal models of β-amyloidopathy (5XFAD) and tauopathy (TauD35) at different ages (Table 3). Considering that PSP is primarily a tauopathy, whereas AD combines features of β-amyloidopathy and tauopathy, we hypothesized that study of these two different animal models could help to uncover pathological processes associated with those hallmarks. Similar to what we observed in the brains of AD patients, the number of DEGs drastically increased with age/pathology progression in both 5XFAD and TauD35 mouse models (Fig 5). Interestingly, however, the number of gDTUs remained low in both models, suggesting that alterations in alternative splicing mechanisms are uncommon in animal models of both β-amyloidopathy and tauopathy. GO analyses revealed that DEGs in 5XFAD mice were enriched for many terms previously observed in the brain of AD patients, such as immune system response, ion homeostasis, response to hormone stimulus, MAPK cascade and synapse signaling (Fig 5A). Also, like AD brains, DEGs were enriched for terms associated with immune-inflammatory response at early pathological stages (4 and 12 months), whereas only at 18 months DEGs were enriched for synapse signaling ontologies. Strikingly, this pattern was upturned in the brains of Taud35 mice (Fig 5B). While at 4 months DEGs/gDTUs were significantly enriched for synapse signaling, only at 17 months they were so for immune system processes.

**Fig 5:**
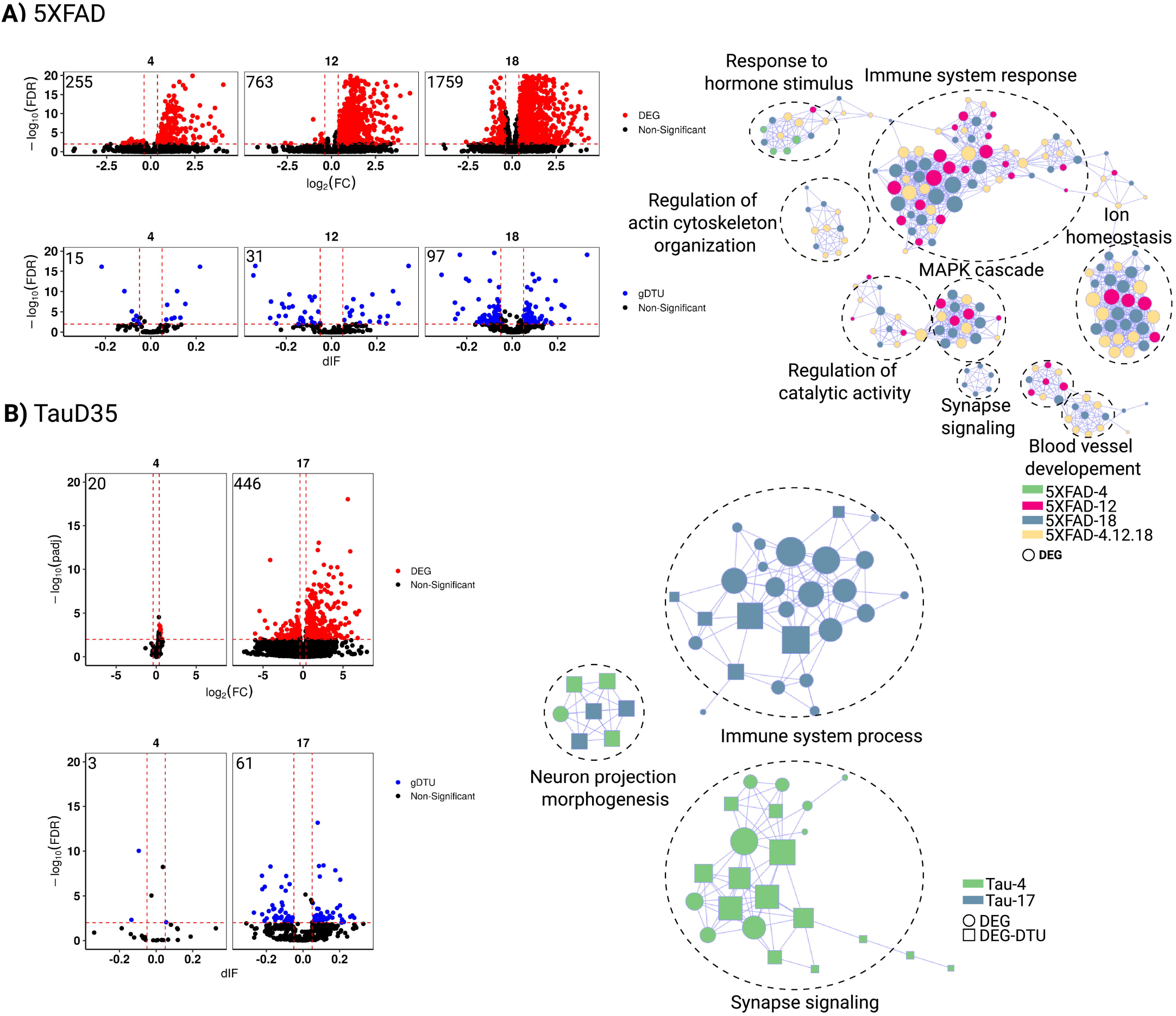
Gene expression alterations in animal models of amyloidopathy and tauopathy. A) Volcano plots showing differentially expressed genes (DEGs, red dots; FC > 1.3 and FDR < 0.01), genes with differential transcript usage (gDTU, blue dots; Differential isoform fraction (dIF) and FDR < 0.01) and a network of ontologies (GOs) for the 5XFAD mouse model data. Circles and squares indicate, respectively, GOs enriched for DEGs or a combination of DEGs and gDTUs. Colors in the network indicate groups where gene expression alterations were detected. B) Same for TauD35 mouse model data. 4 (four months), 12 (twelve months), 17 (seventeen months), 18 (eighteen months). Numbers of significantly altered genes identified in each analysis are shown in the top left corner of the volcano plots

Next, using gene co-expression networks, we identified a module (M1) associated with immune system response in both animal models (Fig 6C) and this module showed higher activity in mutants compared to controls (Fig 6A). Notably, while several DEGs/gDTUs could be detected in the M1 of 5XFAD mice at early stages of pathology (4 months), the vast majority of DEGs in the M1 of Taud35 mice were detected only at late stages (Fig 6B). Conversely, the activity of the modules associated with synapse signaling in 5XFAD mice (M7 and M8) did not show any difference between mutants and controls (Fig 6A). Yet, like human AD brains, DEGs identified in these modules were mainly observed in old animals – 18 months (Fig 6B). Also analogous to human AD and PSP patients, modules associated with extracellular matrix in both animal models (M1_TauD35 and M5_5XFAD) showed higher activity in mutants. On the other hand, we could not detect modules associated with myelination or RNA-splicing in both animal models. Altogether, these findings suggest that β-amyloidopathy primarily leads to an immune-inflammatory response with secondary effects on synapse signaling, whereas tauopathy chiefly affect synapses with subsequent effects in immune-inflammatory activation.

**Fig 6:**
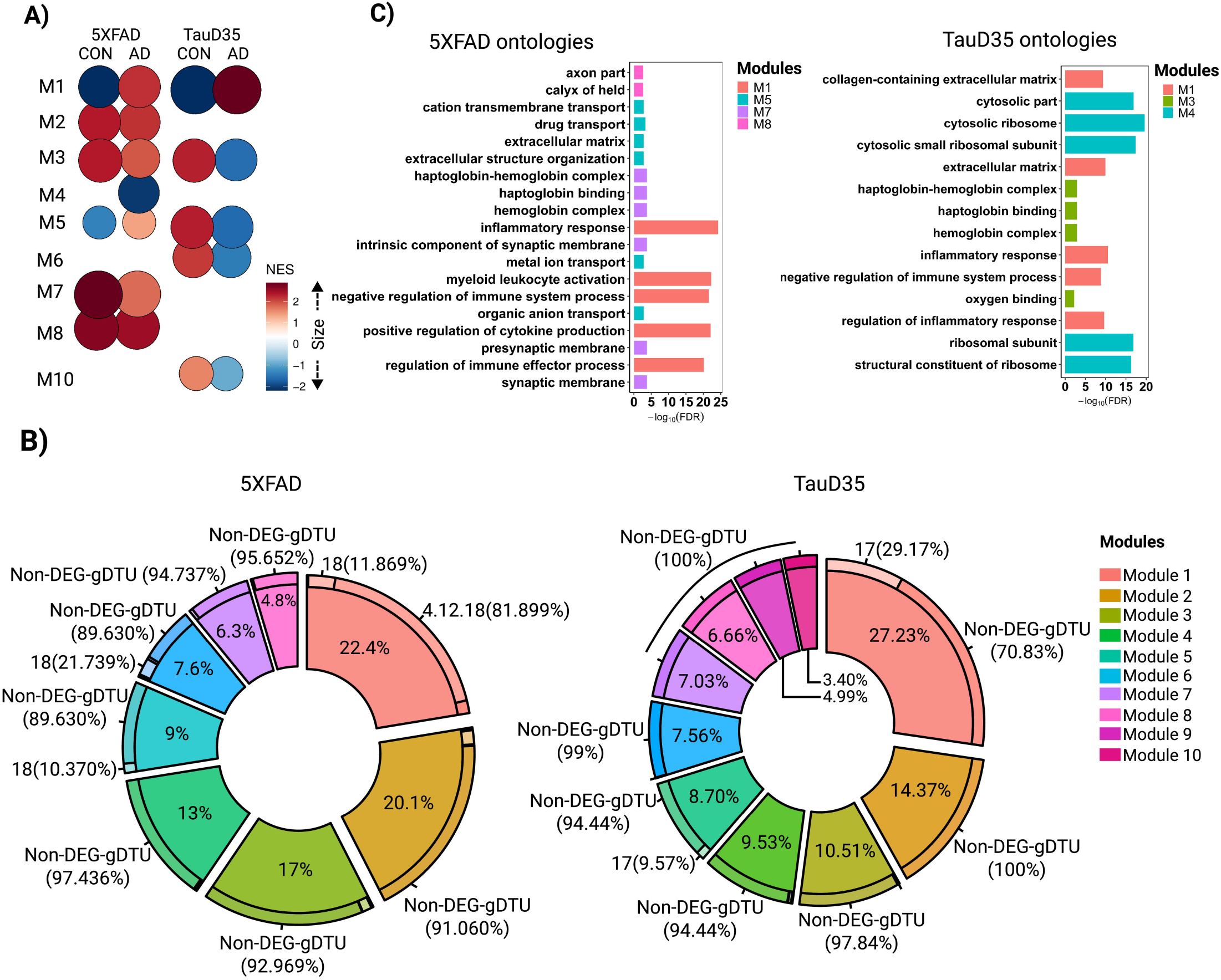
Module enrichment in 5XFAD and TauD35 suggest different time progressions for synaptic alterations and immune-inflammatory responses in these models. A) NES for modules found in 5XFAD and TauD35. Size of circles is equal to the absolute value of NES. Color of circles represents up (red) or down (blue) regulation between classes (Alzheimer and control) and is proportional to NES value. Only modules whose comparison reached statistical significance (FDR<0.01) are shown. B) Quantification of the proportion of genes contributing to each module and the percentages of genes within modules that were DEGs or gDTUs in different groups. C) ORA (Over Representative analysis) form modules found in 5XFAD and TauD35. Only ontologies with FDR < 0.01 are shown.

**Fig 7:**
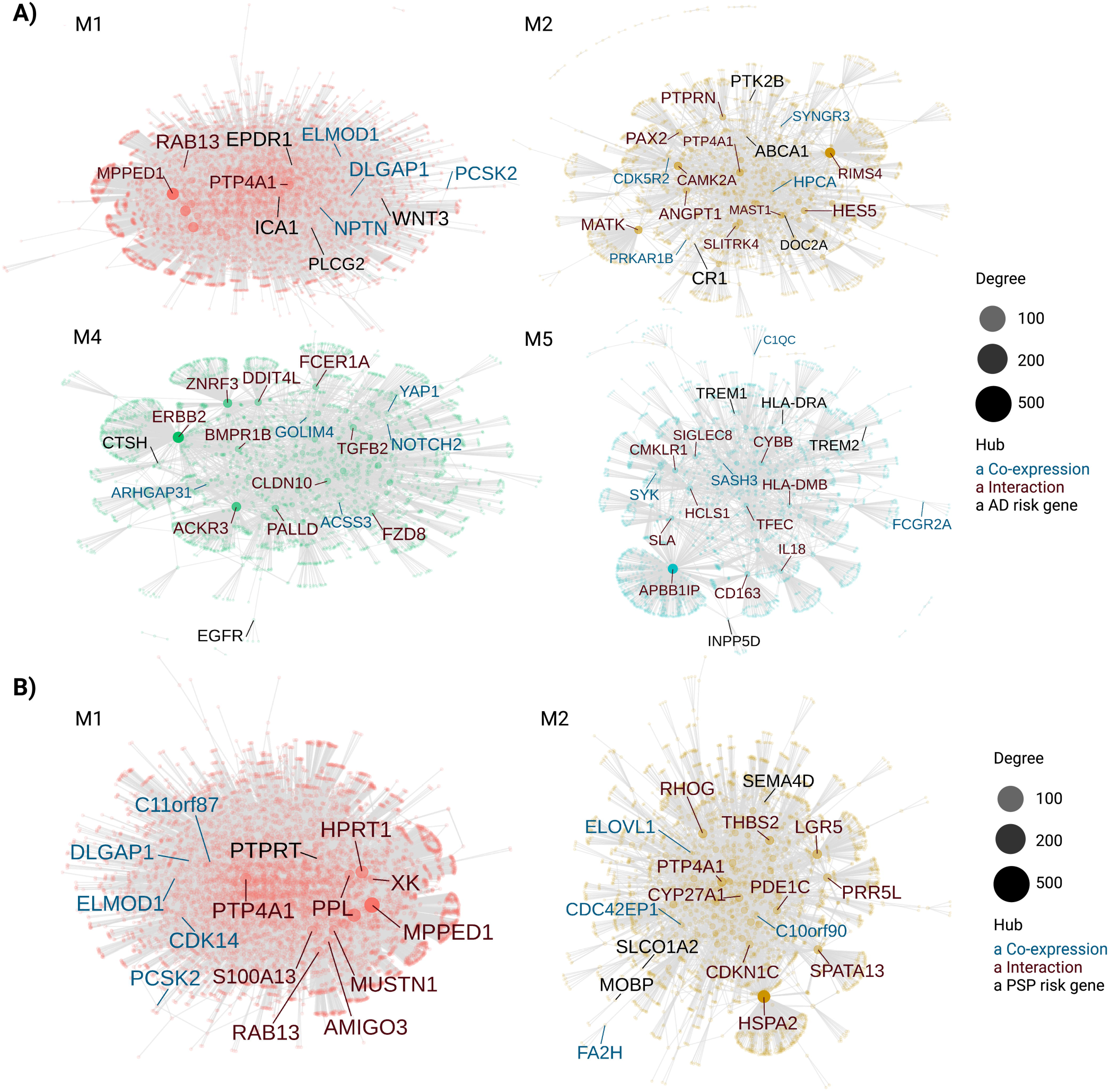
Identification of susceptibility genes for AD and PSP in co-expression modules. A) Protein-protein interaction (PPI) networks for modules containing at least one AD risk genes (black). The five most important hub genes in the PPI based either on co-expression (gene already present in module) or interaction (gene inserted from interactions files of CEMiTool) are shown in blue and red, respectively. Node’s size is proportional to its degree (i.e., number of genes that interacts with a node). B) Same for PSP.

### AD and PSP susceptibility genes are linked to specific gene co-expression networks

Genome-wide association studies (GWAS) have identified polymorphisms in or near several genes that are associated with AD or PSP risk [26, 27, 28]. We used CEMitool to visualize the interactions between these risk genes and the co-expression modules identified in each pathology (Fig 7). In AD modules, we found WNT3 and PLCG2 in M1 (synapse signaling, enriched mainly in glutamatergic neurons), ABCA1, CR1, DOC2A and PTK2B in M2 (synapse signaling, enriched in glutamatergic neurons), CSTH and EGFR in M4 (cellular response to growth factors, enriched in astrocytes), and HLA-DRA, INPP5D, TREM1 and TREM2 in M5 (immune-inflammatory processes, enriched in microglial cells) (Fig 7A). In PSP modules (Fig 7B), we could observe PTPRT in M1 (synapse signaling, enriched in glutamatergic neurons) and MOBP, SEMA4D and SLCO1A2 in M2 (myelination, enriched in oligodendrocytes). Altogether, these results contribute to pinpoint the biological processes likely regulated by 12 AD and 4 PSP genetic risk factors in specific cell types.

## Discussion

Understanding the progression of pathological events in the brain of patients affected by neurodegenerative diseases may help to identify preventive and prognostic-changing treatments for these conditions. In this work, we combined the analysis of transcriptomic data generated from the human brain and a systems biology approach to identify similarities and discrepancies in the biological processes affected in patients diagnosed with two different neurodegenerative diseases. PSP is a primary tauopathy with abnormal accumulation of tau protein within neurons as neurofibrillary tangles (NFTs), primarily in the basal ganglia, diencephalon, brainstem, and cerebellum, with restricted involvement of the neocortex [29]. On the other hand, AD can be considered as a secondary tauopathy, since Aβ plaques are closely tied to the primary neuropathological process. Our findings suggest that, at early ages/stages of disease, tauopathy would be primarily associated with alterations in synaptic signaling processes, whereas amyloidopathy would be mainly associated with immune-inflammatory responses in the brain of PSP and AD patients, respectively. Accordingly, in mouse models of tauopathy and amyloidopathy, gene expression alterations associated with synaptic or immune-inflammatory processes, respectively, predominate at early stages of pathology progression. Last, but not least, we also identify AD risk genes in co-expression modules associated with those biological processes, thus shedding light on their possible contribution to disease onset/progression.

In this study, we analyzed RNAseq data generated from PSP patients showing neuropathological signs of tauopathy in the temporal lobe (Braak stages 1-3, when NFTs are already distinguished the trans-entorhinal and entorhinal cortices) and AD patients with widespread NFTs in the mesocortex, allocortex and neocortex (Braak stages 5-6). Therefore, we believe that the gene expression profile of these samples represents a reasonable proxy of tauopathy- and mixed tauopathy/amyloidopathy-related biological processes altered in the brains of PSP and AD patients, respectively. Additionally, the analysis of RNAseq data obtained from the brains of animal models of tauopathy and amyloidopathy with a well-characterized time progression of pathological processes, allows a more suitable identification of gene expression alterations primarily associated with those processes.

Our observations both in the brains of human patients with PSP and mouse model of tauopathy suggest that biological processes associated with tau accumulation mainly involve neuronal synaptic transmission and that activation of immune-inflammatory processes could be a secondary response. Conversely, in the brains of AD patients and mouse model of amyloidopathy, alterations in gene expression associated with immune-inflammatory response seem to precede or overlap with those related with synapse signaling. These findings are in agreement with previous work using co-expression modules to identify possible intersections between transcriptional alteration in the AD brain and in mouse models of tauopathy or amyloidopathy [13, 10, 7, 8, 9] and may suggest that alterations in synaptic transmission and immune-inflammatory responses are interconnected in a positive feedback loop with different entry points depending on the predominance of tauopathy and amyloidopathy in the brain.

We also show that some biological processes are particularly affected in the brain of AD patients and cannot be fully recapitulated in animal models. This is particularly evident for gene expression alterations associated with RNA splicing processes, which could explain the high number of genes with isoform switches observed in the brains of AD patients [30, 14]. Conversely, gene expression alterations associated with ion homeostasis, response to hormone stimuli, angiogenesis, regulation of protein catabolism, bioenergetics and MAPK cascade were observed both in AD human brains and in 5XFAD mice, but not TauD35 mice, suggesting a link between amyloidopathy and those biological processes. These observations are in accordance with previous work in mice and humans [8, 9, 14] and further support the notion that analysis of gene expression profiles in neurodegenerative diseases is a powerful tool to identify pathology-related alterations.

Our results also show that gene expression alterations associated with myelination, response to growth hormones and angiogenesis could be identified, respectively, in oligodendrocytes, astrocytes and endothelial cells both in AD and PSP brains, suggesting that these biological processes could be common to both pathologies. However, our data indicate that changes in myelination are more prominent in PSP, as it has been previously shown using Weighted Gene Co-Expression Network Analysis [31]. On the other hand, the astrocyte module associated with response to growth factors was significantly affected in AD, but not in PSP brains where we could identify an enrichment for the astrocyte module associated with apoptosis. These alterations could suggest that reactive astrogliosis in AD and PSP are distinct, likely due to the early Tau accumulation in astrocytes observed in the latter [32].

The co-expression of susceptibility genes in cell-type specific modules of AD and PSP revealed in this work is also an interesting hint about the biological processes regulated by those genes. Indeed, we confirm the known roles of INPP5D and TREM2 in the regulation of microglial activation in AD [33, 34] and the involvement of MOBP and SEMA4D in myelinating oligodendrocytes in PSP [31, 35, 36]. Additionally, we provide some interesting hits on the possible contribution of PLCG2, WNT3, ABCA1, CR1 and PTK2B for the regulation of synapse-related processed in glutamatergic neurons, as well as CSTH and EGFR for the regulation of reactive astrogliogenesis in AD. Moreover, we show evidence suggesting that SLCO1A2 and PTRPT contribute to PSP pathogenesis by regulating myelination and synaptic transmission, respectively.

## Conclusions

Altogether, our work provides a comprehensive description of gene expression alterations in the brain of AD and PSP patients, including both gene expression alterations and isoform switches. Using a systems biology approach, we identify pathological processes primarily associated with amyloidopathy and tauopathy in those neurodegenerative diseases, as well as reveal common pathological processes likely resulting from glial and vascular responses. These findings help to open new avenues towards the identification of new pathophysiological processes in AD and PSP, contributing to the development of novel disease-modifying strategies.

## Data Availability

All data referred to in the manuscript is publicly available

## Acknowledgments

The results published here are in whole or in part based on data obtained from the AMP-AD Knowledge Portal (https://adknowledgeportal.synapse.org/). Study data were provided by the following sources: The Mayo Clinic Alzheimer’s Disease Genetic Studies, led by Dr. Nilufer Taner and Dr. Steven G. Younkin, Mayo Clinic, Jacksonville, FL using samples from the Mayo Clinic Study of Aging, the Mayo Clinic Alzheimer’s Disease Research Center, and the Mayo Clinic Brain Bank. Data collection was supported through funding by NIA grants P50 AG016574, R01 AG032990, U01 AG046139, R01 AG018023, U01 AG006576, U01 AG006786, R01 AG025711, R01 AG017216, R01 AG003949, NINDS grant R01 NS080820, CurePSP Foundation, and support from Mayo Foundation. Study data includes samples collected through the Sun Health Research Institute Brain and Body Donation Program of Sun City, Arizona. The Brain and Body Donation Program is supported by the National Institute of Neurological Disorders and Stroke (U24 NS072026 National Brain and Tissue Resource for Parkinson’s Disease and Related Disorders), the National Institute on Aging (P30 AG19610 Arizona Alzheimer’s Disease Core Center), the Arizona Department of Health Services (contract 211002, Arizona Alzheimer’s Research Center), the Arizona Biomedical Research Commission (contracts 4001, 0011, 05-901 and 1001 to the Arizona Parkinson’s Disease Consortium) and the Michael J. Fox Foundation for Parkinson’s Research. Study data were also generated from postmortem brain tissue collected through the Mount Sinai VA Medical Center Brain Bank and were provided by Dr. Eric Schadt from Mount Sinai School of Medicine. We also thank GSK and Company scientists for generating the TauD35 RNASeq data and providing us access to them. 5XFAD mice are provided through the IU/JAX/UCI MODEL-AD Center, established with funding from The National Institute on Aging (U54 AG054345-01 and AG054349). Aging studies are also supported by the Nathan Shock Center of Excellence in the Basic Biology of Aging (NIH P30 AG0380770).

## Supporting information captions

Fig S1: Unique gene expression alterations in temporal cortex and cerebellum at different ages. A) DEGs identified in temporal cortex data of AD and PSP. B) Intercept graphic showing the overlap among DEGs and gDTUS identified in the temporal cortex of AD and PSP patients compared to controls. C) Same as in (A) for cerebellum. D) Same as in (B) for cerebellum. Up (genes with higher expression when compared to control individuals), Down (genes with lower expression when compared to control individuals).

Fig S2: Gene expression alterations in the cerebellum of AD and PSP patients. A) Volcano plots showing differentially expressed genes (DEGs, red dots; FC > 1.3 and FDR < 0.01), genes with differential transcript usage (gDTU, blue dots; Differential isoform fraction (dIF) and FDR < 0.01) and a network representation of gene ontologies (GOs) significantly enriched in AD. Circles, triangles and squares indicate, respectively, GOs enriched for DEGs, gDTUs or a combination of both. Colors in the network indicate groups where gene expression alterations were detected. B) Same for PSP. AD (Alzheimer), PSP (Progressive Supranuclear Palsy), A (age of death between 70-80 years old), B (age of death between 81-89 years old), C (age of death equal or superior to 90 years old), FDR (False Discovery Rate). Numbers of significantly altered genes identified in each analysis are shown in the top left corner of the volcano plots.

Fig S3: Gene expression alterations associated with immune system and synapses follow pathology progression in AD brains. A) NES for modules identified in the PRh and RLPFC of AD patients compared to controls. Size of circles is equal to the absolute value of NES. Color of circles represents up (red) or down (blue) regulation between classes (AD and control) and is proportional to NES value. B-C) Over Representative Analysis (ORA) of RLPFC and PRh modules. Only ontologies with FDR < 0.01 are shown.

Fig S4: Isoform switches (gDTUS) are rare in the brains of AD animal models. A-B) DEGs identified in the brains of 5XFAD (A) and TauD35 (B) compared to control animals. C) Intercept graphic showing the overlap among DEGs identified in the models. Up (genes with higher expression when compared to control animals), Down (genes with lower expression when compared to control animals).

## Description of supplementary tables

Table S1: DEGs and gDTUs identified in the brain of AD or PSP patients compared to controls, and in the brain of 5XFAD and TauD35 mutants compared to wild type mice. Data are organized by group/age.

Table S2: Gene ontologies significantly enriched after inputting DEGs, gDTUs or both in gprofiler2’s R package from human and animal models data analyzed. Data are organized by group/age.

Table S3: Over-representation analysis of all co-expression modules identified in human and animal models data.

Table S4: List of genes in the different co-expression modules identified by CEMiTool in the human and mouse brains.

