## Supplementary figures and images for "Analysis of modular gene co-expression networks reveals molecular pathways underlying Alzheimer’s disease and progressive supranuclear palsy"

### Figure S1

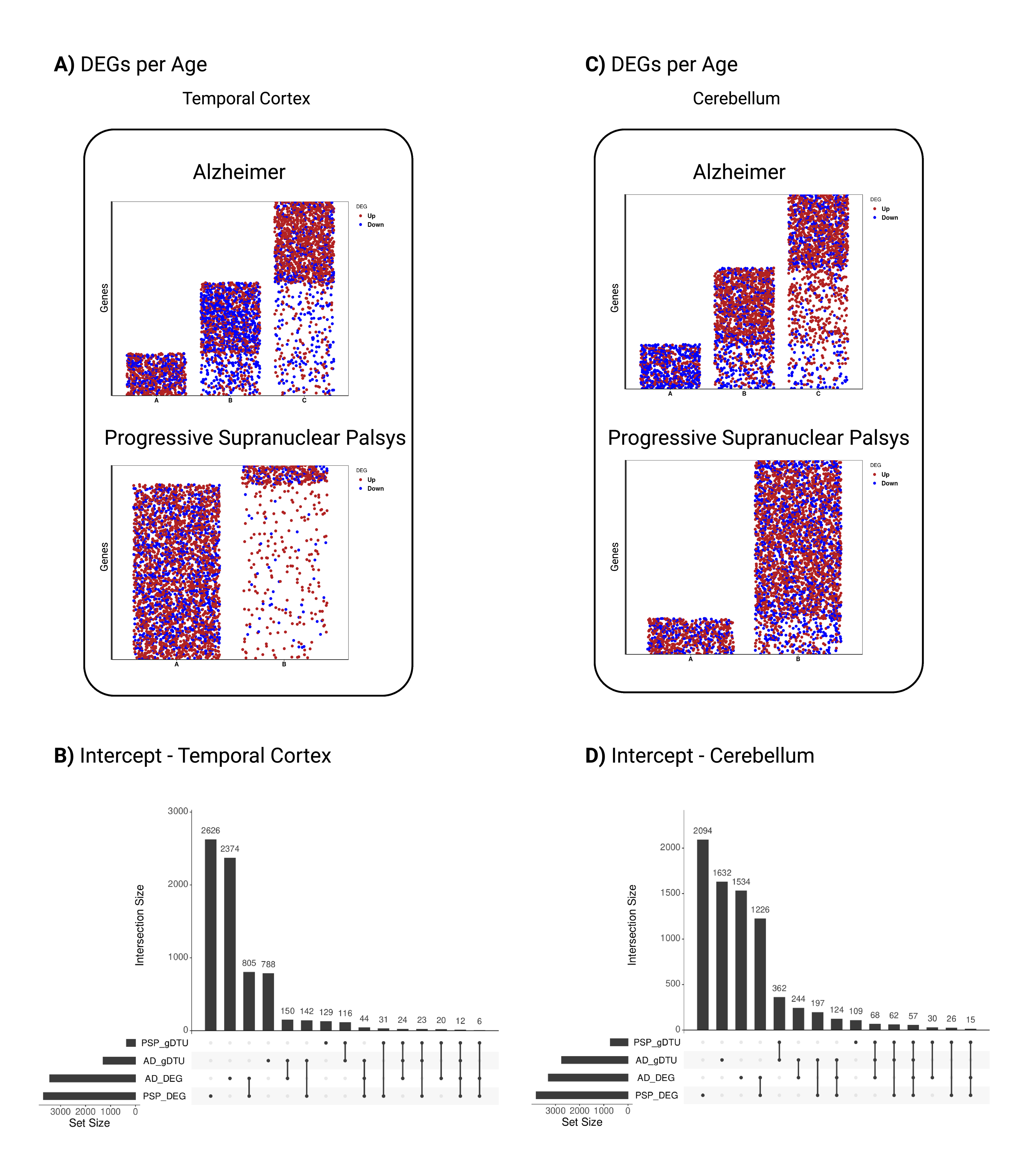

### Figure S2

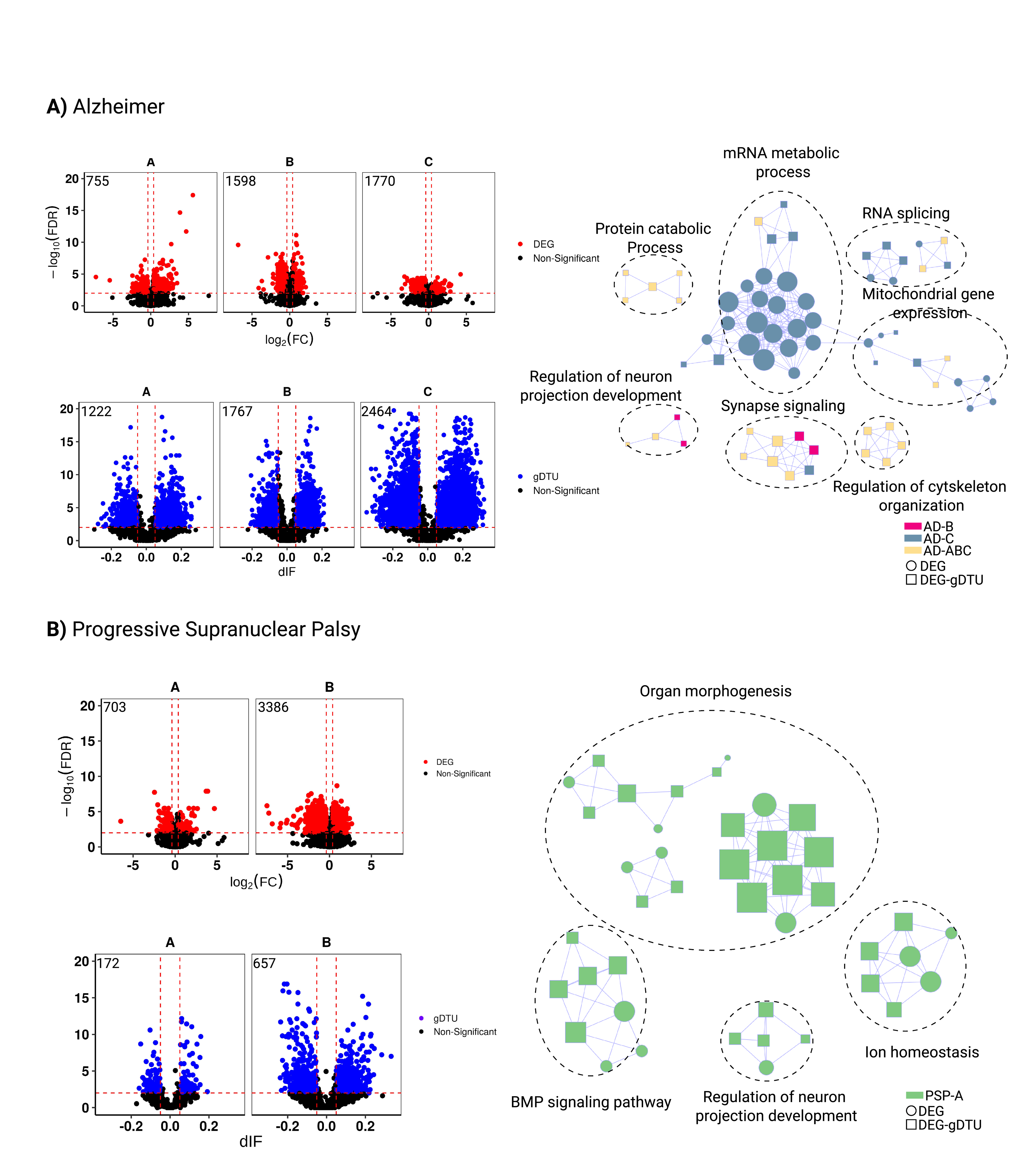

### Figure S3

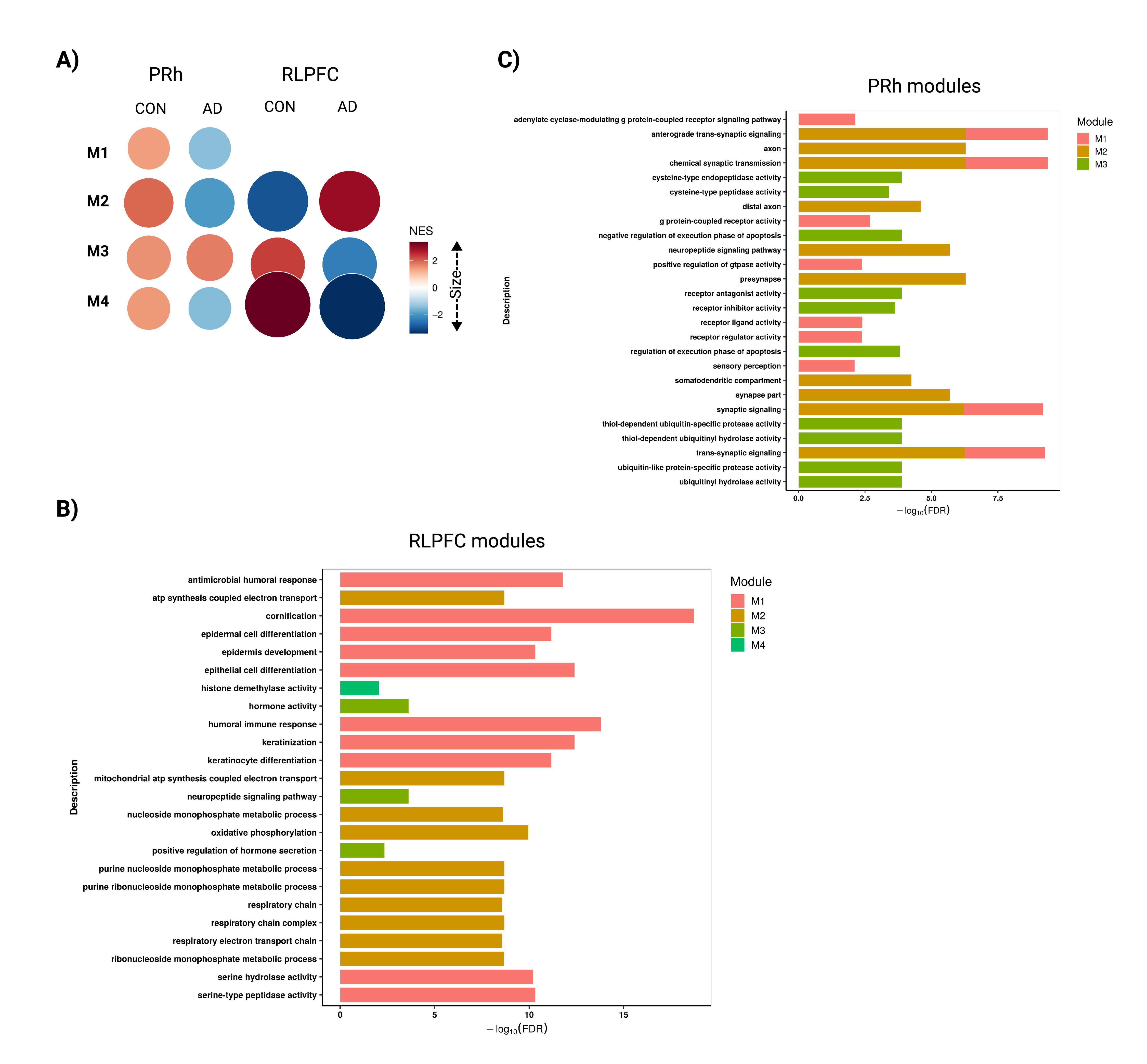

### Figure S4

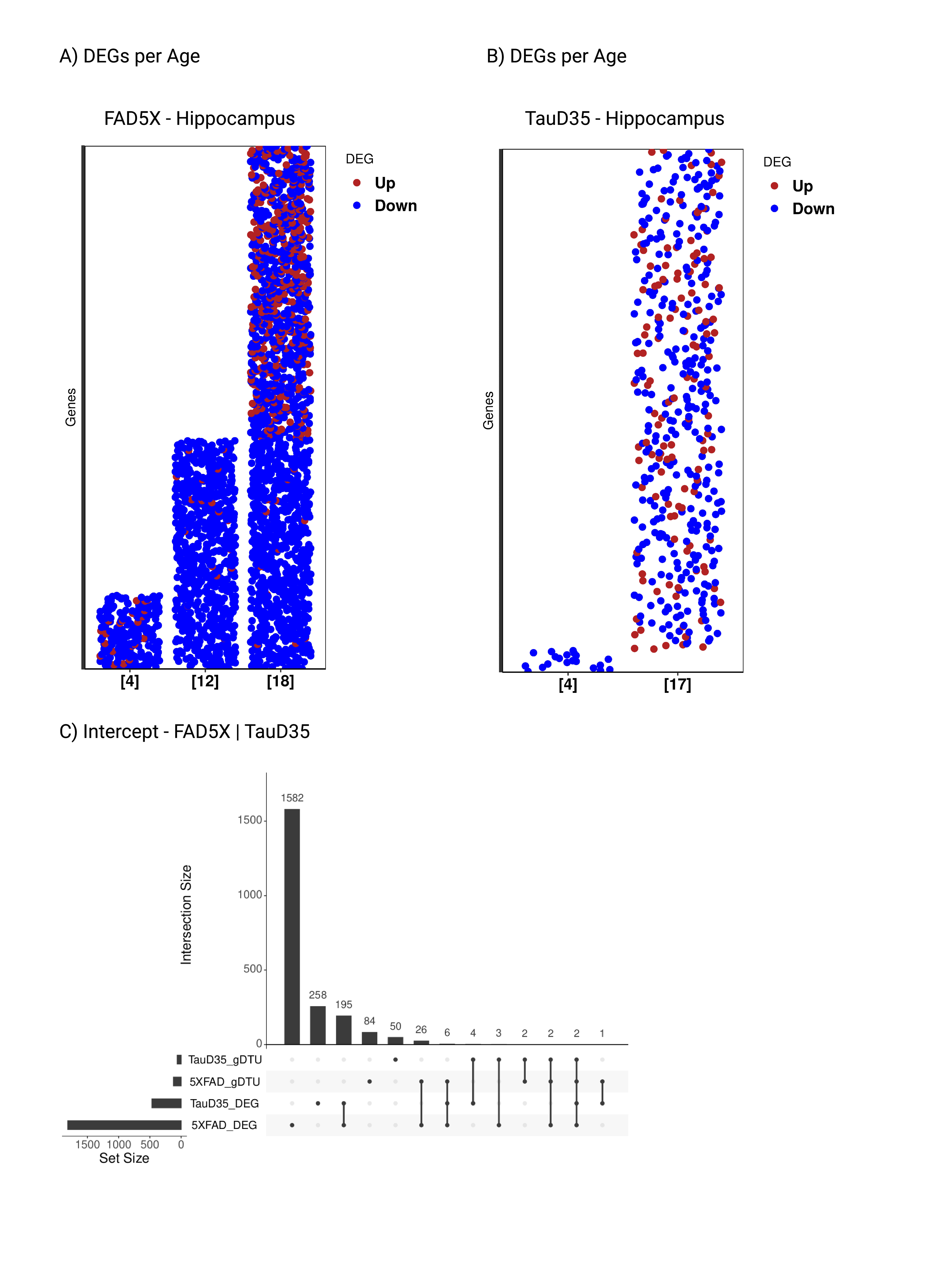
